# Evaluation of the OPTN Six-Status Heart Allocation System and a Combined Prognostic Model for Waitlist Risk Evaluation

**DOI:** 10.1101/2025.09.22.25336410

**Authors:** Elizabeth J. Bashian, Michael T. Cain, Bryon Bhagwandin, Bruce Kaplan, Jordan R.H. Hoffman, John S. Malamon

## Abstract

**KEY POINTS:** *Question:* Does the current national six-status heart allocation system rank waitlisted transplant patients based on medical urgency? Can this heart allocation system’s prognostic performance be improved?

*Findings:* The six-status system is poorly calibrated, lacks sufficient statistical discrimination, and underestimates risk in the highest-risk patients, or those with an observed six-month mortality probability greater than 2%. By combining the current status system with six additional patient characteristics (previous transplant, ventilation, mean pulmonary capillary wedge pressure, willingness to accept a donor after cardiac death, diabetes status, and most recent creatinine), we correctly predicted greater than 80% of six-month waitlist mortalities in 7,706 study participants.

*Meaning:* This study challenges the safety and efficacy of the current national heart allocation system.

**Importance:** In December 2016, the Organ Procurement and Transplantation Network (OPTN) approved a bylaw that restructured the national heart allocation policy from a three-status to a six-status system. This new allocation system, which aimed to assign the highest priority to the patients with the highest mortality risk, went into effect on October 18, 2018. Since then, studies have identified limitations with the current system. However, no changes have been made to improve the national heart allocation system.

**Objective:** Given the clear importance and impact of ranking patients based on medical urgency, we carefully evaluated the six-status heart allocation system to determine its correlation with observed mortality, or calibration, and its ability to predict six-month patient mortality risk and waitlist survival. We identified six additional patient characteristics associated with waitlist mortality and combined them with the six-status score to significantly improve the current allocation system’s ability to predict six-month waitlist mortality.

**Design:** A retrospective, secondary analysis of the Scientific Registry of Transplant Recipients (SRTR) database of heart transplant candidates and recipients waitlisted from October 18, 2018, to December 31, 2024.

**Setting:** The United States

**Participants:** Single-organ heart transplant candidates, 18 years of age and older who were placed on the waitlist (N = 19,275). Patients listed multi-organ transplantation were excluded.

**Exposures:** All-cause waitlist mortality

**Main Outcomes and Measures:** The primary outcome of this study was the validation of the calibration and prognostic performance of the current heart allocation system. The secondary outcome is a simple model that greatly improves upon the current system’s ability to accurately (>80%) predict waitlisted patient mortality.

**Results:** With a mean calibration slope of 0.94 (0.66, 1.21) and an area under the receiver operating curve of 0.71 (0.47, 0.87), the current allocation system is poorly calibrated, has only moderate statistical discrimination, and underestimates patient risk in the most critically ill patients. Hazard and time-series regression analysis confirmed that the six-status system does not adequately rank patients based on medical urgency. Our combined model demonstrates that the national allocation system can be improved.

**Conclusions and Relevance:** While the current heart distribution system accounts for some patient risk factors, a more objective and accurate model is needed to achieve the OPTN’s strategic objective to more reliably model and predict patient risk and survival likelihood. Our model more accurately predicts patient waitlist mortality and will better inform waitlist management and improve waitlist survival by prioritizing medical urgency.

## 1. INTRODUCTION

Since 1988, the Organ Procurement and Transplantation Network (OPTN) has been engaged in the development and refinement of a cardiac transplant allocation system that allocates donated hearts to the most severely ill patients^1^. On October 18, 2018, a revised heart allocation policy and bylaw were implemented, transitioning from a 3-tiered to a 6-tiered status-based system utilizing a patient risk stratification model designed to address the rapidly increasing complexity of managing critical cardiac patients through medical judgment and exception criteria^2^. The goal of the new allocation system was to reduce waitlist mortality by better stratifying the high-risk patient groups and to improve equity in transplant allocation. The six-status system has been criticized for its subjectivity and variability in effectively distinguishing true patient mortality risk while on the waitlist. For example, a large proportion of transplant candidates are now stratified as Status 1 or Status 2 by exception rather than by the standard criteria, which can be modified by a physician’s practice^3^. Early studies on post-transplant survival have demonstrated that the 2018 allocation policy revision was associated with a significant reduction in post-transplant survival^4,5^. However, in 2022, Lazenby *et al.* performed a time-dependent survival analysis to demonstrate the effect of inadequate follow-up on post-policy survival results and their interpretation. They concluded that there was no significant difference in 1-year post-transplant patient survival under the new heart allocation policy, despite the goal of the new system being to provide organs to patients most likely to survive^6^. Other studies have replicated these findings^7^. However, the six-status system has been criticized for its subjectivity and variability in effectively distinguishing true patient mortality risk while on the waitlist^5,8,9^.

For example, a large proportion of transplant candidates are now stratified as Status 1 or Status 2 by exception rather than by the standard criteria, which can be modified by a physician’s practice^3^. In addition, it has been shown that baseline status at time of listing does not accurately predict waitlist mortality^9^. Similarly, Pelzer et al. determined that the current allocation system had only a moderate ability to successfully rank transplant candidates according to medical urgency^10^. Therefore, a formal prognostic model that reliably stratifies and ranks waitlisted end-stage heart failure (HF) patients according to medical urgency is urgently needed to provide impartiality, precision, and transparency to better inform decision-making and contextualize the complex relationship between patient risk and benefit.

The current six-status system ranks patients from Status 1 to 6, where Status 1 is the highest priority. Status-1 patients must have received extracorporeal membrane oxygenation (ECMO), a nondischargeable surgically implanted ventricular assist device, or a durable LVAD with life-threatening arrhythmias. Status-2 patients must have received a durable LVAD with an intra-aortic balloon pump (IABP), percutaneous endovascular LVAD, surgically implanted non-endovascular LVAD, discharged with a total artificial heart, or sustained ventricular arrhythmias. Status-3 patients must have received durable LVAD or inotropic agents with continuous hemodynamic monitoring.

Status-4 patients have durable LVAD without complications, inotropic agents without continuous hemodynamic monitoring, cardiomyopathies, intractable angina, or were listed for re-transplantation. Status-5 patients are dual-organ transplants, and Status-6 patients include all other waitlisted candidates.

Predictive models that integrate patient history with serology have been successfully implemented in other organ transplant systems. The Model for End-stage Liver Disease, including sodium, or MELD-Na, score for liver transplantation accurately predicts 90-day patient waitlist mortality and is the most significant metric in liver allocation. A similar model is desperately needed in cardiac transplantation to accurately identify and prioritize the most critically ill patients. The primary aim of this study is to utilize the Scientific Registry of Transplant Recipients (SRTR) database to evaluate the six-status heart allocation system and determine its correlation with observed mortality and ability to predict six-month patient mortality. Finally, we combined the current six-status system with six additional patient variables to develop a logistic regression model which marks a significant improvement in the predictive performance using the current allocation system.

## 2. METHODS

### 2.1 Data sources

This study utilized retrospective data from the SRTR. The SRTR system includes data on all donors, waitlisted candidates, and transplant recipients in the US, submitted by the members of the OPTN^12^. The Health Resources and Services Administration, U.S. Department of Health and Human Services provides oversight to the activities of the OPTN and SRTR contractors. The data reported here have been supplied by the Hennepin Healthcare Research Institute as the contractor for SRTR. The interpretation and reporting of these data are the responsibility of the author(s) and in no way should be seen as an official policy of or interpretation by the SRTR or the U.S. Government.

### 2.2 Study population

The study population included adult waitlisted cardiac transplant patients (N=19,264) who registered and were listed for a heart transplant between October 18, 2018, and December 31, 2024. All study participants were retrospectively censored at two time points, first on October 18, 2018, and then on March 2, 2025, to provide a full six months of patient follow-up days. (**Supplemental Figure 1).** Survival times for waitlisted candidates started at the date of listing and were censored at the date of death or removal from the waitlist. Patients removed from the waitlist for all-cause mortality were censored. All patients were censored at the date of the last administrative follow-up, June 2, 2025. This population was randomly split (60% by 40%) into two cohorts: a training set (N= 11,558) and a test set (N=7,706). **Table 1** provides the population characteristics of the two randomly selected study groups. The training set was used to train each model, and the test set was used to independently evaluate the six-status system’s performance in predicting mortality outcomes and to rank patients’ relative risk of death after six months on the waitlist. The training set was used to generate logistic regression equations, and the test set was used to independently evaluate model performance.

**Table 1.**
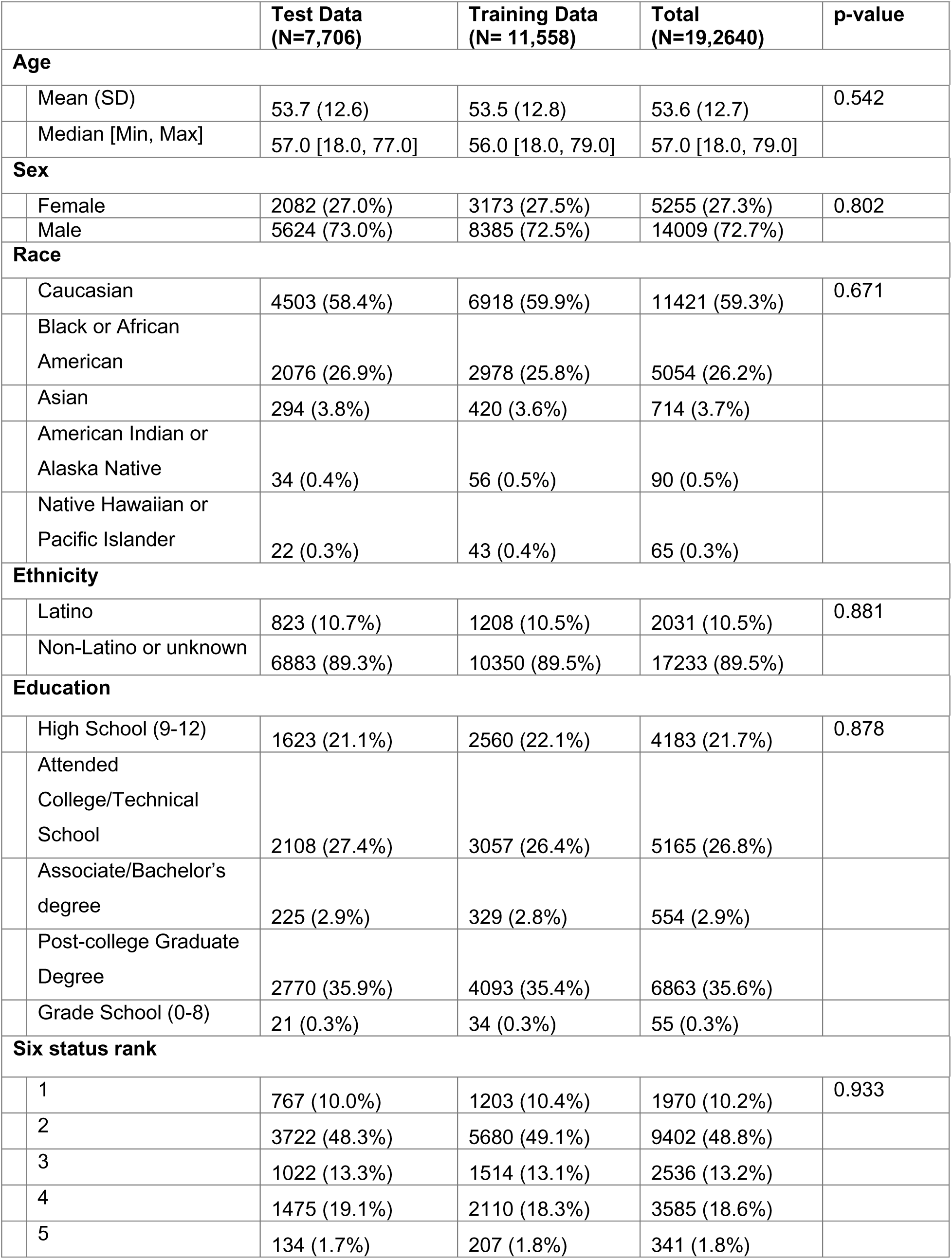
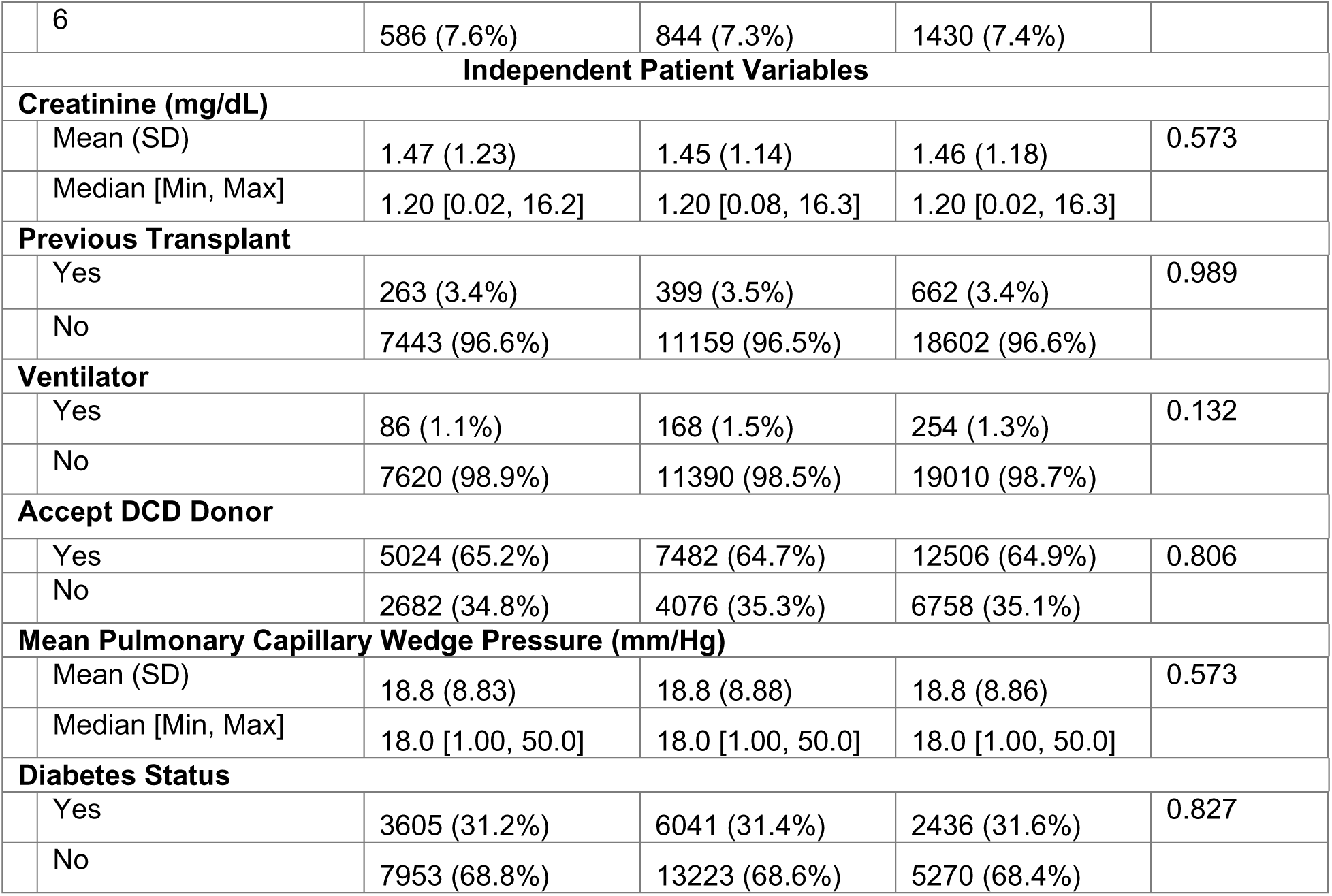
Patient Characteristics of Heart Transplant Patients Waitlisted from 2018 to 2024.

|  | Test Data<br>(N=7,706) | Training Data<br>(N= 11,558) | Total<br>(N=19,2640) | p-value |
| --- | --- | --- | --- | --- |
| **Age** |  |  |  |  |
| Mean (SD) | 53.7 (12.6) | 53.5 (12.8) | 53.6 (12.7) | 0.542 |
| Median [Min, Max] | 57.0 [18.0, 77.0] | 56.0 [18.0, 79.0] | 57.0 [18.0, 79.0] |  |
| **Sex** |  |  |  |  |
| Female | 2082 (27.0%) | 3173 (27.5%) | 5255 (27.3%) | 0.802 |
| Male | 5624 (73.0%) | 8385 (72.5%) | 14009 (72.7%) |  |
| **Race** |  |  |  |  |
| Caucasian | 4503 (58.4%) | 6918 (59.9%) | 11421 (59.3%) | 0.671 |
| Black or African<br>American | 2076 (26.9%) | 2978 (25.8%) | 5054 (26.2%) |  |
| Asian | 294 (3.8%) | 420 (3.6%) | 714 (3.7%) |  |
| American Indian or<br>Alaska Native | 34 (0.4%) | 56 (0.5%) | 90 (0.5%) |  |
| Native Hawaiian or<br>Pacific Islander | 22 (0.3%) | 43 (0.4%) | 65 (0.3%) |  |
| **Ethnicity** |  |  |  |  |
| Latino | 823 (10.7%) | 1208 (10.5%) | 2031 (10.5%) | 0.881 |
| Non-Latino or unknown | 6883 (89.3%) | 10350 (89.5%) | 17233 (89.5%) |  |
| **Education** |  |  |  |  |
| High School (9-12) | 1623 (21.1%) | 2560 (22.1%) | 4183 (21.7%) | 0.878 |
| Attended<br>College/Technical<br>School | 2108 (27.4%) | 3057 (26.4%) | 5165 (26.8%) |  |
| Associate/Bachelor's<br>degree | 225 (2.9%) | 329 (2.8%) | 554 (2.9%) |  |
| Post-college Graduate<br>Degree | 2770 (35.9%) | 4093 (35.4%) | 6863 (35.6%) |  |
| Grade School (0-8) | 21 (0.3%) | 34 (0.3%) | 55 (0.3%) |  |
| **Six status rank** |  |  |  |  |
| 1 | 767 (10.0%) | 1203 (10.4%) | 1970 (10.2%) | 0.933 |
| 2 | 3722 (48.3%) | 5680 (49.1%) | 9402 (48.8%) |  |
| 3 | 1022 (13.3%) | 1514 (13.1%) | 2536 (13.2%) |  |
| 4 | 1475 (19.1%) | 2110 (18.3%) | 3585 (18.6%) |  |
| 5 | 134 (1.7%) | 207 (1.8%) | 341 (1.8%) |  |
| 6 | 586 (7.6%) | 844 (7.3%) | 1430 (7.4%) |  |
| **Independent Patient Variables** |  |  |  |  |
| **Creatinine (mg/dL)** |  |  |  |  |
| Mean (SD) | 1.47 (1.23) | 1.45 (1.14) | 1.46 (1.18) | 0.573 |
| Median [Min, Max] | 1.20 [0.02, 16.2] | 1.20 [0.08, 16.3] | 1.20 [0.02, 16.3] |  |
| **Previous Transplant** |  |  |  |  |
| Yes | 263 (3.4%) | 399 (3.5%) | 662 (3.4%) | 0.989 |
| No | 7443 (96.6%) | 11159 (96.5%) | 18602 (96.6%) |  |
| **Ventilator** |  |  |  |  |
| Yes | 86 (1.1%) | 168 (1.5%) | 254 (1.3%) | 0.132 |
| No | 7620 (98.9%) | 11390 (98.5%) | 19010 (98.7%) |  |
| **Accept DCD Donor** |  |  |  |  |
| Yes | 5024 (65.2%) | 7482 (64.7%) | 12506 (64.9%) | 0.806 |
| No | 2682 (34.8%) | 4076 (35.3%) | 6758 (35.1%) |  |
| **Mean Pulmonary Capillary Wedge Pressure (mm/Hg)** |  |  |  |  |
| Mean (SD) | 18.8 (8.83) | 18.8 (8.88) | 18.8 (8.86) | 0.573 |
| Median [Min, Max] | 18.0 [1.00, 50.0] | 18.0 [1.00, 50.0] | 18.0 [1.00, 50.0] |  |
| **Diabetes Status** |  |  |  |  |
| Yes | 3605 (31.2%) | 6041 (31.4%) | 2436 (31.6%) | 0.827 |
| No | 7953 (68.8%) | 13223 (68.6%) | 5270 (68.4%) |  |

### 2.3 Study Ethics

Informed consent was obtained for all study participants. This study was reviewed by an ethical committee (Colorado Multiple Institutions Review Board) and was determined to be non-human subjects research. This study was not grant funded.

The authors do not have any conflicts of interest to disclose. All authors reviewed the results and approved the final manuscript.

### 2.4 Statistical Approach

#### 2.4.1. Summary Statistics

For continuous variables, a two-way ANOVA test was used to test the observed differences in patient characteristics and the independent variables used in this study. The chi-squared test was used to measure the significance of categorical and indicator (binary) variables. Patient predictor variables should exhibit discrimination and low collinearity. To measure collinearity, we calculated Pearson’s cross-correlation coefficient (R) and variance inflation factor (VIF) for all independent variables. Low collinearity was defined as two independent variables with an R value less than 0.7. The VIF is used to determine the correlation between independent variables in a logistic regression model. A VIF of 1 provides no correlation, whereas values above 2.5 indicate considerable multicollinearity^13^.

#### 2.4.2. Binned Residuals

Binned residuals were calculated and plotted using R’s ‘performance’ package. Binned residuals are the differences between the observed mortality and the OPTN six-status allocation system, where bins represent each risk stratum, or status level, ranging from 1 to 6. This method was used to determine the goodness-of-fit between true and predicted patient morality^14–16^.

#### 2.4.3 Model Calibration

A well-calibrated risk ranking system is required for the accurate estimation of patient mortality risk, or medical urgency. To assess the calibration of the six-status system, we utilized R’s ‘CalibrationCurves’ package^17,18^. Using a generalized linear model with a logit link function, this software package provided the calibration intercept, slope, and discrimination (c-statistic). ‘CalibrationCurves’ works by describing the correlation of risk predictions by using calibration curves. A calibration curve maps the predicted probabilities *f*(***x***_*i*_) to the actual event probabilities *P*(*y*_*i*_=1|*f*(***x***_*i*_)), where ***x***_*i*_was the six-status rank index and *y*_*i*_ was a patient death. This provided a visualization of the correspondence between the model’s predicted and observed patient mortality probabilities^17–19^. All analyses were performed using the R statistical language version 4.4.3. The TRIPOD protocol, or the transparent reporting of a multivariable prediction model for individual prognosis or diagnosis, was followed throughout this study^20^.

#### 2.4.4 Validating Dichotomous Outcomes in the Combined Model

Because the six-status score provides some statistical discrimination, we hypothesized that we could leverage this information in combination with other common patient characteristics to improve prediction performance. By adding, previous transplant (yes/no), ventilation (yes/no), mean pulmonary capillary wedge pressure, willingness to accept a donor after cardiac death (DCD), diabetes status (yes/no), and most recent creatinine. We measured this model’s performance in predicting mortality events using sample hold-out validation. We calculated coefficients using the training set, and we measured the AUC and other performance metrics using the test or validation set. To avoid confirmation bias in selecting the training and test sets, we performed 30 random splits and recorded the AUC of each test. R’s ‘pROC’ package was used to calculate the 95% confidence intervals of each model. All patient risk scores were calculated using **Equations 1-4**. Coefficients were calculated using the discovery set, and predictions were independently validated using the validation set. Finally, we calculated the area under the ROC curve, or AUC.

## 3. RESULTS

### 3.1 Population Characteristics

The training and test patient population characteristics are depicted in **Table 1**. The mean (SD) age of the 19,264 study participants ranged from 18 to 79 years of age, with a mean age of 53.59 (12.69) years. 72.7% of participants were male, 68.4% were White or Caucasian, 26.2% were Black or African American, 3.7% were Asian, 0.34% were Native Hawaiian or Other Pacific Islander, and 0.47% were American Indian or Alaska Native. In summary, there were no statistically significant differences in the patient characteristics or independent variables when comparing the discovery set to the validation set. Importantly, there were no significant differences in the p-values of the training and test sets.

### 3.2 Binned Residuals and Calibration Curves

**Figure 1A** provides the average residual as a function of the estimated patient mortality probability for each risk stratum, represented by the black dots. The average residual is the mean of the differences between the observed six-month mortality probability and the predicted mortality probability within each of the six status groups. Estimated six-month mortality probabilities ranged from 0.2 to 5%. Importantly, all six status tiers (labeled in red) had statistically significantly negative residuals, demonstrating that the estimated mortality probability is lower than the expected probability. Thus, the current six-status system significantly underestimates patient risk. Also, the status tiers are not ordinally ranked with regard to the observed mortality rate. **Figure 1B** demonstrates the calibration curve and discrimination performance results. The calibration intercept and slope were 0.20 (−0.4, 0.44) and 0.88 (0.58-1.18), respectively. The c-statistic was 0.69 (0.48-0.84). Importantly, we can see that the six-status system was severely miscalibrated for the highest-risk patients, or those with a six-month mortality rate greater than 3%.

**Figure 1.**
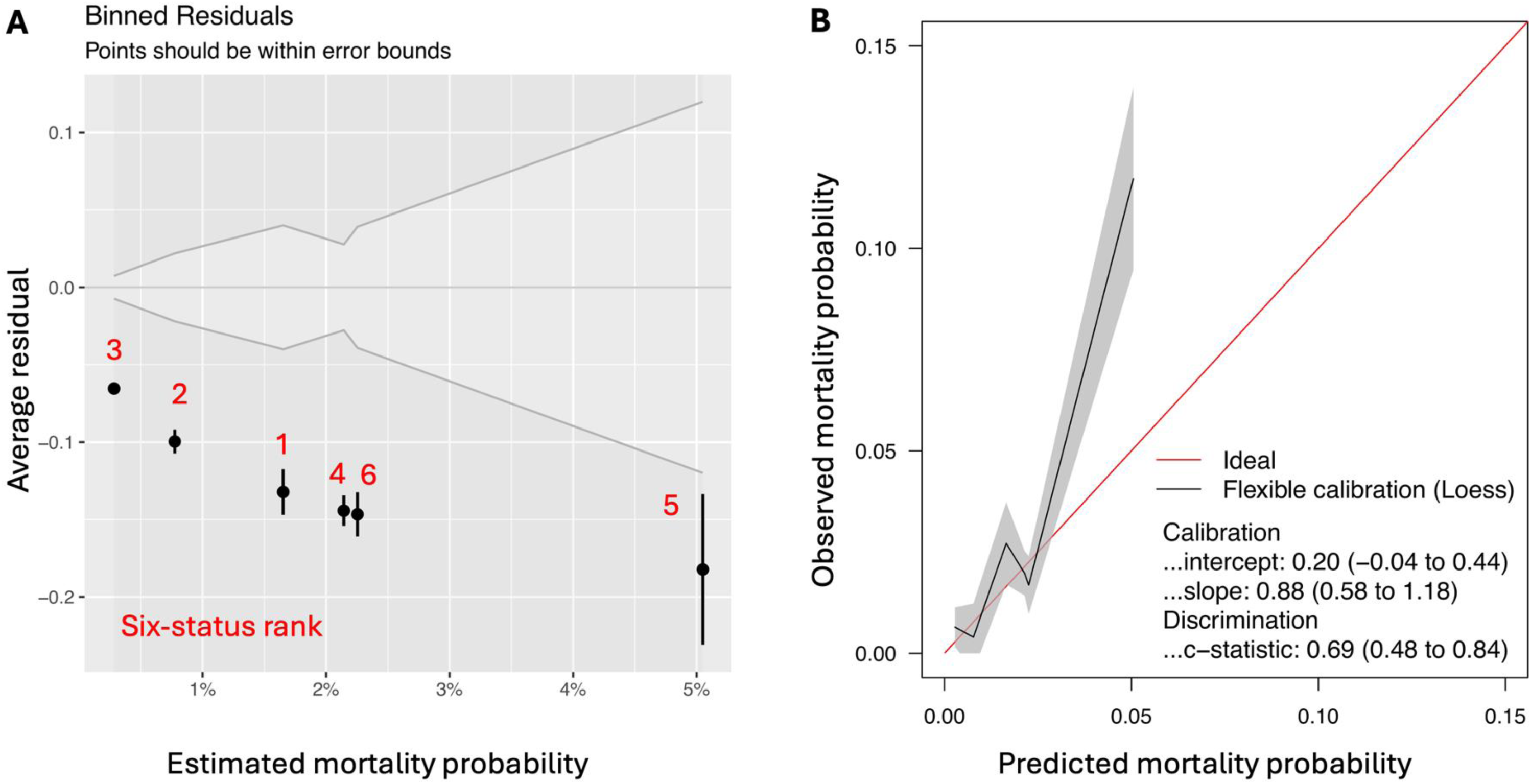
Binned residuals and calibration plots. Figure 1A provides the average residual as a function of the estimated patient mortality probability for each risk stratum, represented by the black dots. The average residual is the mean of the differences between the observed six-month mortality probability and the predicted mortality probability within each of the six status groups. Estimated six-month mortality probabilities ranged from 0.2 to 5%. Importantly, all six status tiers (labeled in red) had statistically significantly negative residuals, demonstrating that the estimated mortality probability is lower than the expected probability. Figure 1B demonstrates the calibration curve and discrimination performance results. The calibration intercept and slope were 0.20 (−0.4, 0.44) and 0.88 (0.58-1.18), respectively. The c-statistic was 0.69 (0.48-0.84).

### 3.3 Cox Proportional-hazard Hazard Regression Analysis

We applied the Cox proportional-hazard model as a univariate and time-dependent validation test, with the six-tiered rank as a predictor. **Figure 2A** provides the six-month hazard ratios and 95% confidence intervals for all tiers relative to Status 1, or the reference stratum. Status 2 and 3 had significantly lower mortality risk (0.34 and 0.35, *p*<0.008) than Status 1, as expected; however, the two strata are not significantly different in terms of six-month waitlist survival. Interestingly, Status 5 was by far the highest risk status, with a relative hazard ratio of 5.57 (2.97, 10.43, *p*<0.001).

**Figure 2.**
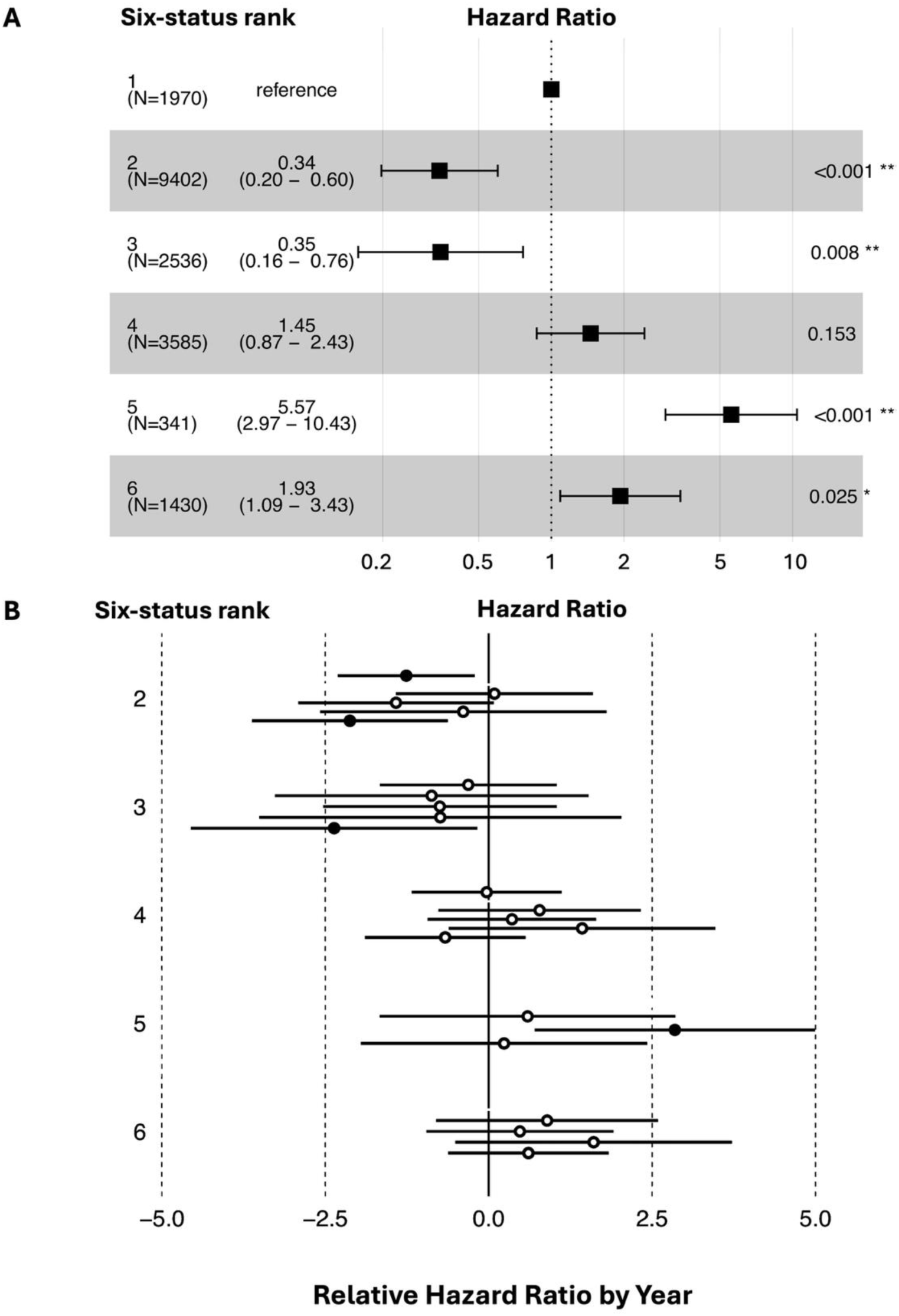
Cox proportional-hazard ratio of the six-status system. Figure 2A provides the six-month hazard ratios and 95% confidence intervals for all tiers relative to Status 1, or the reference stratum. Status 2 and 3 had significantly lower mortality risk (0.34 and 0.35, *p*<0.008) than Status 1, as expected; however, the two strata are not significantly different in terms of six-month waitlist survival. Interestingly, Status 5 was by far the highest risk status, with a relative hazard ratio of 5.57 (2.97, 10.43, *p*<0.001). Surprisingly, Status 6 was associated with significantly higher mortality risk than Status 1, with a hazard ratio of 1.93 (1.09, 3.43, *p*=0.025). Figure 2B provides the relative hazard ratio for each tier relative to Status 1, where each horizontal bar is the relative Cox HR estimation for a full calendar year, ranging from 2019 to 2024.

Surprisingly, Status 6 was associated with significantly higher mortality risk than Status 1, with a hazard ratio of 1.93 (1.09, 3.43, *p*=0.025). **Figure 2B** provides the relative hazard ratio for each tier relative to Status 1, where each horizontal bar is the relative Cox HR estimation for a full calendar year, ranging from 2019 to 2024. Status 2 and 3 are similarly somewhat reduced but not always significantly different, and the HR for Status 5 is significantly greater than Status 1. This analysis also highlights the significant heterogeneity, and year-to-year variability observed in the current allocation system.

### 3.4 Kaplan-Meier Survival Analysis

Six-month survival analysis was performed for each status level. **Figure 3** provided Kaplan-Meier survival probability curves and risk tables for all six levels. Survival analysis confirmed CPHR analysis, where it can be observed that Status-5 patients have the lowest survival probability (p<0.0001). Similarly, Status-2 and Status-3 patients have the highest survival probability (greater than 99.5%, p<0.0001).

**Figure 3.**
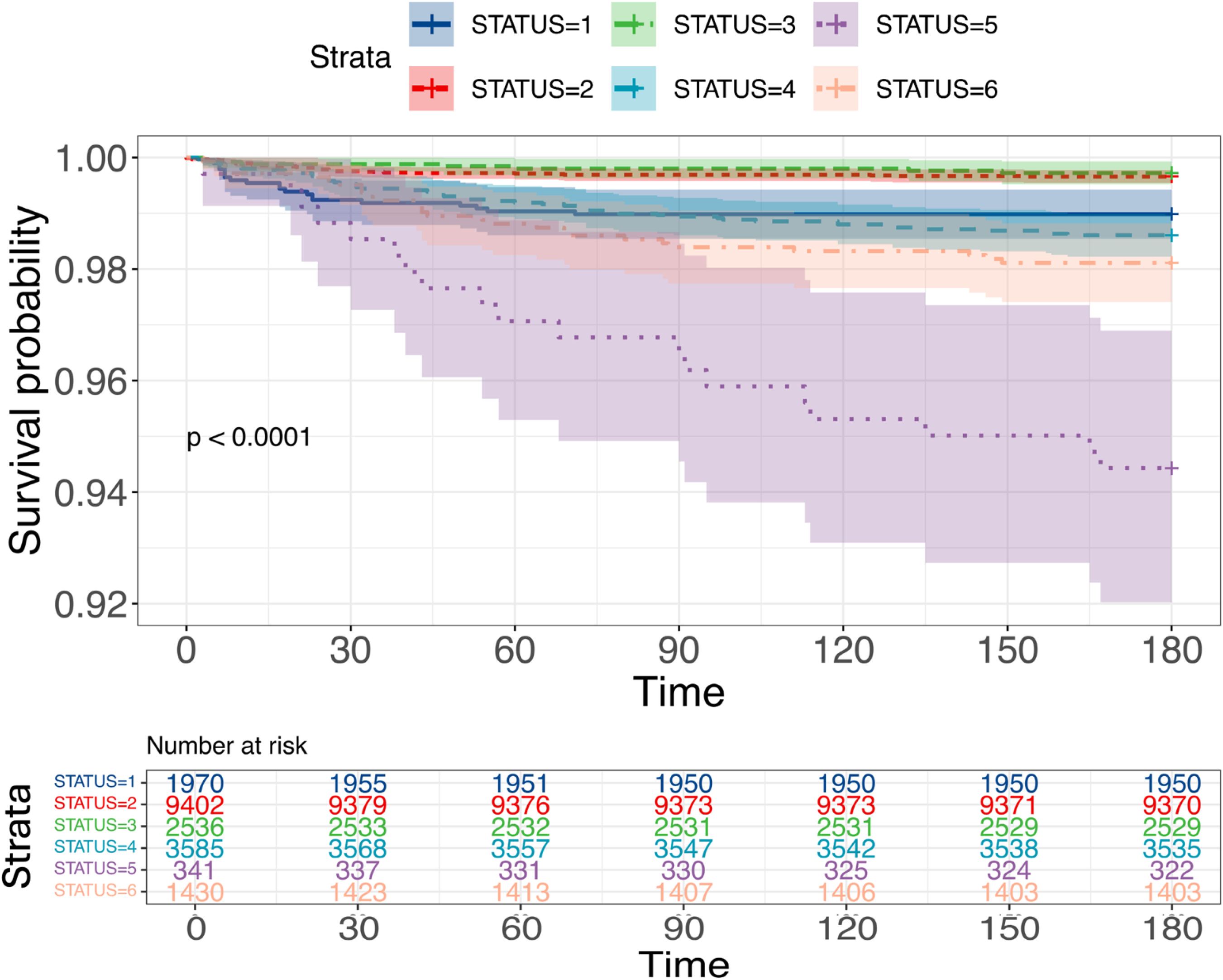
**Six-month Kaplan-Meier survival probability curves of the six-status system.** Kaplan-Meier survival probability curves are provided for each of the six status tiers. Risk tables are also provided for each status in the table below.

### 3.5 Logistic Regression Classification Performance and Formula

Using independent sample hold-out validation, the calculated AUC (mean/SD) was 0.8089 (0.7522-0.8656, **Figure 4**). All thirty random validation sets based on selection splits fell within the 95% confidence intervals. In summary, this model provides a more accurate methodology for predicting the six-month waitlist survival likelihood of waitlisted heart transplant patients. **Eq. 1** provides the logistic regression formula for each of the six-status strata and six patient characteristics. **Supplemental Table 1** contains odds ratios, standard errors, confidence intervals, and p-values for all independent variables used in the combined model.

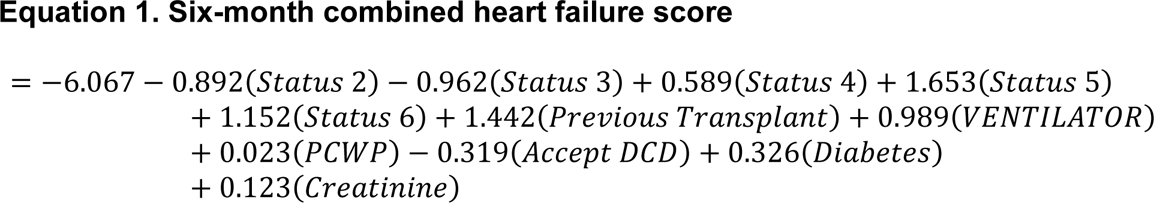

**Figure 4.**
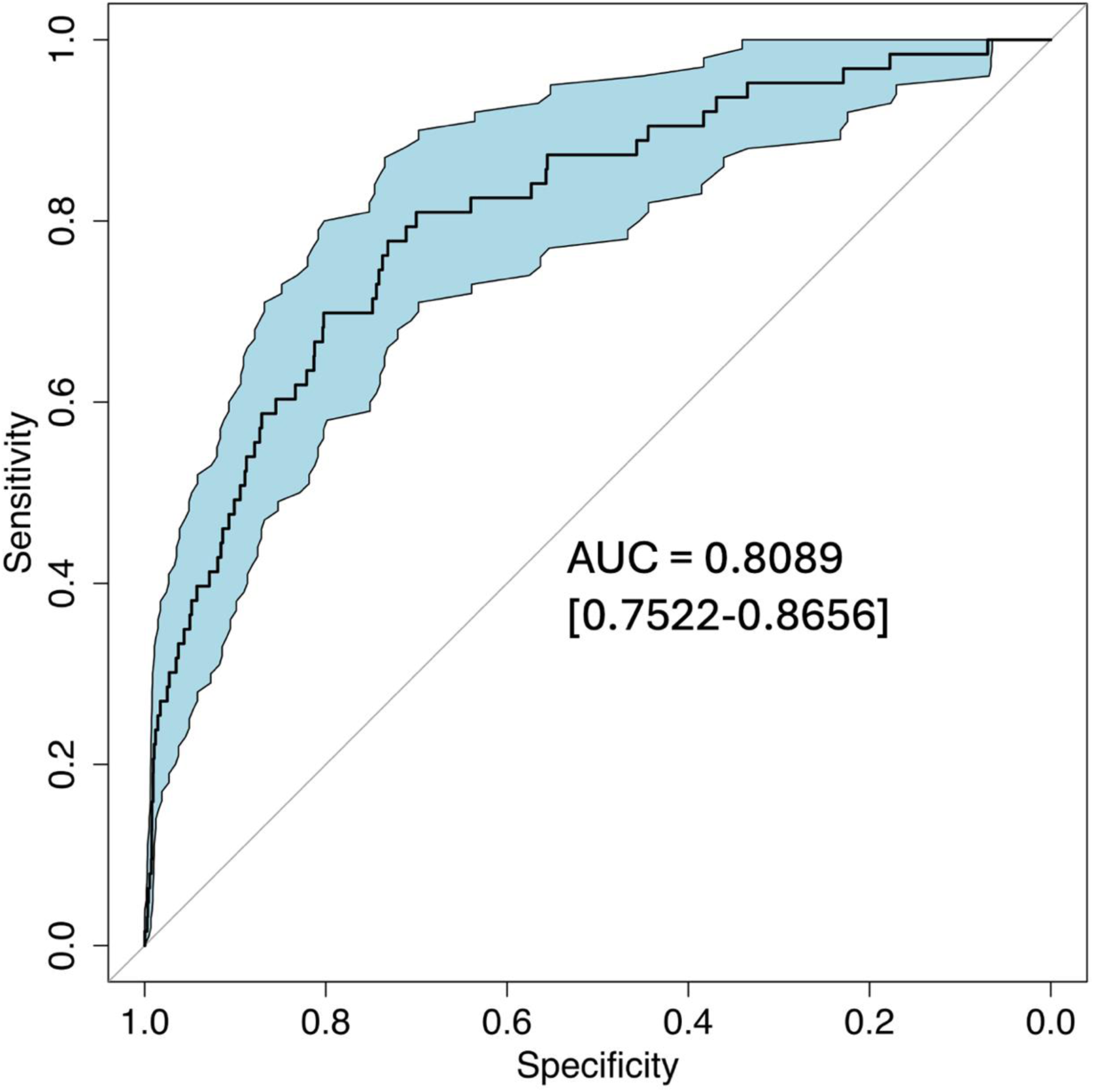
**Area under the receiver operating curve of the combined model.** We constructed a logistic regression model using the six-status rank and six additional patient characteristics: creatinine, previous transplant, use of ventilation, willingness to accept a DCD donor, mean pulmonary capillary wedge pressure, and diabetes status. Using independent sample hold-out validation, the calculated area under the receiver operating curve (mean/CI) was 0.8089 (0.7522-0.8656). The model was trained with the training set (N=11,558), and validation was performed using the test set (N=7,706).

## 4. DISCUSSION

This study evaluated the calibration and performance of the current six-status allocation system’s performance in predicting six-month waitlist mortality and proposed an augmented model incorporating six additional clinical variables to greatly improve upon the current system. In 2023, it was demonstrated that the OPTN’s six-status heart allocation system had only a moderate ability (AUC=0.67) to identify the short-term survival likelihood of waitlisted heart transplant candidates^10^. Our analysis of the six-status system confirmed that the current allocation system only moderately predicts waitlist outcomes, with an average AUC of 0.69. Importantly, calibration analysis revealed that the current allocation system does not reliably stratify patients by medical urgency but, in fact, significantly underestimates waitlist mortality probability in patients with a six-month waitlist mortality rate above 3%. Significant year-to-year heterogeneity was observed, showing significant variation in the six-status system’s performance over time and possibly explicating the seemingly contradictory pre- and post-transplant survival analysis results under this organ allocation system.

While useful prognostic tools for patients with HF have been introduced, such as the Heart Failure Survival Score^21^ and the Seattle Heart Failure Model^22^, they have not accurately predicted patient waitlist mortality^23^. The Ottawa Heart Failure Risk Scale provided a risk stratification tool designed for acute heart failure patients in emergency departments^24^. However, this score has not been applied to or validated against a waitlisted heart transplant patient population. Similarly, the Meta-Analysis Global Group in Chronic Heart Failure (MAGGIC) risk score performed well for HF patients with preserved ejection fraction, but it has not been externally validated for reduced ejection fraction, which excludes most patients on a heart transplant waitlist^25^. More recently, the US-CRS model highlighted the need for an accurate and objective waitlist mortality model^26^. Finally, we introduced the Colorado Heart failure Acuity Risk Model, or CHARM score, which used six patient serology metrics and seven medical history indicator variables to accurately (AUC > 0.8) predict short-term patient morality risk^27^.

Predictive models that integrate patient history with serology have been successfully implemented in other organ transplant systems. The Model for End-stage Liver Disease, including sodium, or MELD-Na, score for liver transplantation accurately predicts 90-day patient waitlist mortality and is the most significant metric in liver allocation^28^. For example, Evans *et al*. demonstrated an overall 1-year survival rate increase of 18% in high-acuity patients in the 15 years following the national implementation of the MELD-Na score and then MELD 3.0.^29^ Similarly, the Acute Physiology and Chronic Health Evaluation (APACHE) score integrates laboratory tests, comorbidities, and patient vital signs to assist with the appropriate management of critically ill patients^30^. A similar model is desperately needed in cardiac transplantation to accurately identify and prioritize the most critically ill patients.

Because the six-status system does provide some statistical discrimination, for practicality, we choose to combine it with six additional patient variables to more accurately predict six-month waitlist survival likelihood. These six variables have been previously used to predict waitlist survival, are consistently reported in the SRTR, and improve statistical discrimination. These included previous transplant status, ventilator dependency, mean PCWP, willingness to accept a DCD donor, diabetes status, and most recent serum creatinine, which were chosen because they are associated with a higher risk of waitlist mortality. By adding these variables to the existing allocation system, we achieved an average AUC of 0.8089, demonstrating improved statistical discrimination. This marks an absolute increase of 10% in correctly ranking patient mortality likelihood. This model has the ability to significantly improve waitlist survival.

While the OPTN’s current system for allocating heart transplants has improved the equity of distribution of donor organs with no impact on patient outcomes, these improvements have come at the expense of some undesirable events, such as longer travel times to donor hospitals and longer donor allograft ischemic times^7,31,32^.

Additionally, advances in mechanical circulatory support and other bridge-to-transplant interventions have significantly impacted waitlist survival^4^. Bridging interventions such as the implantation of a temporary LVAD or other circulatory support devices have significantly reduced patient waitlist risk and mortality^33,34^. At the same time, durable LVAD patients are given lower priority, which is associated with increased waitlist complications^7,33,34^. A similar effect is seen with patients needing retransplantation. In addition, the widespread use of exceptions further undermines the current allocation system’s objectivity. A recent analysis showed that 15-18 percent of patients are upgraded to a higher status by exception. Although listing by exception does not improve overall survival after transplant, waitlist survival rates are significantly higher than those of standard candidates^35^. Exception criteria is based on approval by regional boards and lacks uniformity; however, 95% of exception requests are approved nationwide^35^. This is especially notable given that the revised allocation system was created to decrease the number of exceptions and has instead had the opposite effect. Exception requests were more commonly seen at centers that had high-status 1A volume under the old allocation system, suggesting strategic behavior to maintain volume^36^. An evidence-based, data-driven prognostic model could better support allocation decisions by reducing subjectivity and improving transparency. Because the SRTR does not record exceptions in its thoracic candidate waitlist tables, we were not able to examine their effects on waitlist mortality, a limitation of this study.

The implications relating to policy development are significant, as an allocation system more reflective of medical urgency may help reduce avoidable waitlist mortality and improve overall outcomes while continuing to improve access to organ allocation. The experience of other organ systems, such as the MELD-Na score in liver transplantation, supports the potential for data-driven scoring systems to improve equity and outcomes. While the current system accounts for many patient factors, an objective model that can accurately stratify patients by medical urgency is needed to better inform patients of their true risk and the trade-off between risk and benefit. Formal pre-transplant patient mortality models have been successfully developed and utilized in heart, liver, and kidney transplantation and have resulted in reduced patient mortality. A similar metric is needed for cardiac transplantation to identify and prioritize the most critically ill waitlisted patients.

In conclusion, the current OPTN six-status heart allocation system does not reliably stratify patients by medical urgency and significantly underestimates mortality risk in the most critically ill candidates. Our findings demonstrate that incorporating additional objective clinical variables markedly improved the prediction of the six-month waitlist mortality. The revised model offers a more accurate and equitable approach to ranking transplant candidates. Given the limitations of the exception-based prioritization, a data-driven allocation system is urgently needed. Adoption of a formal risk model could improve waitlist survival and support more consistent, transparent organ distribution. These results provide a compelling foundation for future revisions to the heart allocation policy.

### Author Contributions

JSM and BB had full access to all the data in the study and take responsibility for the integrity of the data and the accuracy of the data analysis.

*Concept and design: JSM, JRHH, and MTC*

*Acquisition and analysis of data: BB and JSM*

*Interpretation of data:* All authors.

*Drafting of the manuscript: EB, MTC, JRHH, JSM*

*Critical revision of the manuscript:* All authors.

*Statistical analysis: JSM and BB*

*Obtained funding: BK*

*Administrative, technical, or material support: BB, EB, and JSM*

*Supervision: BK, JRHH, JSM, MTC*

## CONFLICTS OF INTEREST

The authors do not have any conflicts of interest to disclose.

## Data Availability

All data are publically available

## ACKNOWLEDGMENT

The authors have no acknowledgment to make.

## ABBREVIATIONS

AUC: Area under the curve
ANOVA: Analysis of variance
CI: Confidence interval
CHARM: Colorado Heart Failure Acuity Risk Model
CPHR: Cox proportional-hazard regression
ECMO: Extracorporeal membrane oxygenation
HF: Heart failure
HR: Hazard ratio
IABP: Intra-aortic balloon pump
ICD: Implantable cardiac defibrillators
MAGGIC: Meta-Analysis global group in chronic heart failure
MELD: The model for end-stage liver disease
OPTN: Organ procurement and transplantation network
PCWP: Pulmonary capillary wedge pressure
ROC: Receiver operating characteristic
SRTR: Scientific registry of transplant recipients
SD: Standard deviation
LVAD: Left ventricular assist devices

---

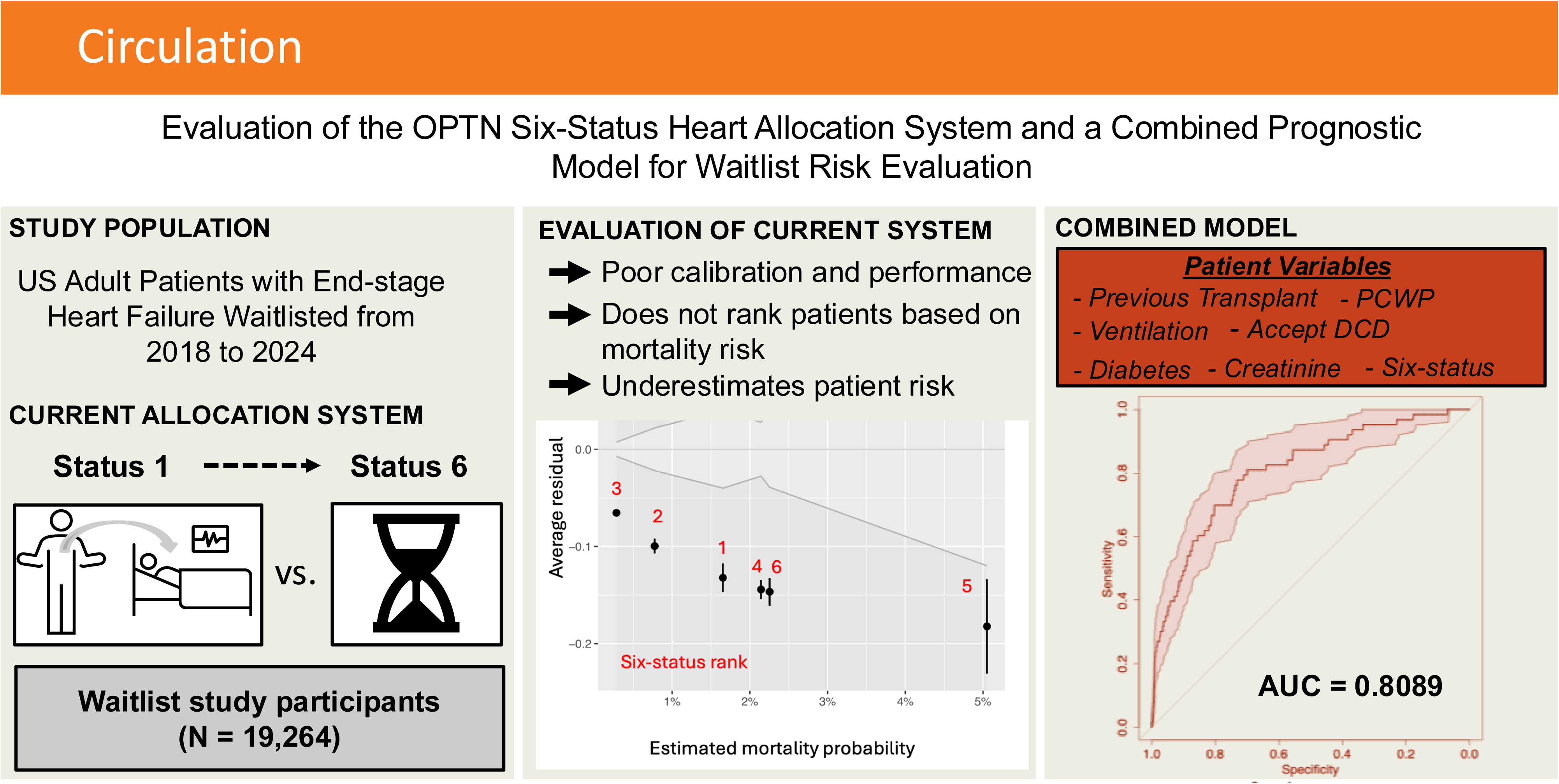

## Notes

### Competing Interest Statement

The authors have declared no competing interest.

### Funding Statement

No funding was using in this research

### Author Declarations

The Colorado Multiple Institutional Review Board (COMIRB) approved all research

## REFERENCES

1. Stevenson LW. Crisis Awaiting Heart Transplantation: Sinking the Lifeboat. JAMA Intern Med. 2015;175(8):1406–1409. doi:10.1001/jamainternmed.2015.2203

2. Adult heart allocation - OPTN. Accessed October 20, 2024. https://optn.transplant.hrsa.gov/professionals/by-organ/heart-lung/adult-heart-allocation/

3. Maitra NS, Dugger SJ, Balachandran IC, Civitello AB, Khazanie P, Rogers JG. Impact of the 2018 UNOS Heart Transplant Policy Changes on Patient Outcomes. JACC Heart Fail. 2023;11(5):491-503. doi:10.1016/j.jchf.2023.01.009

4. Cogswell R, John R, Estep JD, et al. An early investigation of outcomes with the new 2018 donor heart allocation system in the United States. J Heart Lung Transplant. 2020;39(1):1–4. doi:10.1016/j.healun.2019.11.002

5. Kilic A, Mathier MA, Hickey GW, et al. Evolving Trends in Adult Heart Transplant With the 2018 Heart Allocation Policy Change. JAMA Cardiol. 2021;6(2):159–167. doi:10.1001/jamacardio.2020.4909

6. Lazenby KA, Narang N, Pelzer KM, Ran G, Parker WF. An updated estimate of posttransplant survival after implementation of the new donor heart allocation policy. Am J Transplant. 2022;22(6):1683–1690. doi:10.1111/ajt.16931

7. Estep JD, Soltesz E, Cogswell R. The new heart transplant allocation system: Early observations and mechanical circulatory support considerations. J Thorac Cardiovasc Surg. Published online September 16, 2020:S0022–5223(20)32638-6. doi:10.1016/j.jtcvs.2020.08.113

8. Patel JN, Abramov D, Fudim M, Okwuosa IS, Rabkin DG, Chung JS. The heart transplant allocation change attenuates but does not eliminate blood group O waitlist outcome disadvantage. Clin Transplant. 2022;36(5):e14620. doi:10.1111/ctr.14620

9. Blackstone EH, Rajeswaran J, Cruz VB, et al. Continuously Updated Estimation of Heart Transplant Waitlist Mortality. J Am Coll Cardiol. 2018;72(6):650–659. doi:10.1016/j.jacc.2018.05.045

10. Pelzer KM, Zhang KC, Lazenby KA, et al. The Accuracy of Initial U.S. Heart Transplant Candidate Rankings. JACC Heart Fail. 2023;11(5):504–512. doi:10.1016/j.jchf.2023.02.005

11. Alam A, Hall S. Policy and Oversight of Cardiac Transplantation. Methodist DeBakey Cardiovasc J. 21(3):83–91. doi:10.14797/mdcvj.1567

12. Learn about EPTS - OPTN. Accessed June 22, 2025. https://optn.transplant.hrsa.gov/data/allocation-calculators/epts-calculator/learn-about-epts/

13. Johnston R, Jones K, Manley D. Confounding and collinearity in regression analysis: a cautionary tale and an alternative procedure, illustrated by studies of British voting behaviour. Qual Quant. 2018;52(4):1957–1976. doi:10.1007/s11135-017-0584-6

14. tutorial on calibration measurements and calibration models for clinical prediction models | Journal of the American Medical Informatics Association | Oxford Academic. Accessed June 22, 2025. https://academic.oup.com/jamia/article/27/4/621/5762806

15. Residuals and Diagnostics for Ordinal Regression Models: A Surrogate Approach: Journal of the American Statistical Association: Vol 113, No 522. Accessed June 22, 2025. https://www.tandfonline.com/doi/abs/10.1080/01621459.2017.1292915?casa_token=XApXeInxCSUAAAAA:9M0NdxnJFNYbLMCG5cMhgpcnYQf5Uevt1MhlFanZG2wH_MLhlTCoegmLebLm97aPJyc9OLOXDJo

16. Modeling continuous response variables using ordinal regression - Liu - 2017 - Statistics in Medicine - Wiley Online Library. Accessed June 22, 2025. https://onlinelibrary.wiley.com/doi/abs/10.1002/sim.7433?casa_token=dDJKzUKJZP4AAAAA%3AsQswJG7E-_bm-7UvAj0cQU4qbLt0lacefjGhnpZx6kzQmzJNRiC7NxmCuzNYoepshZaf60ntZUya

17. Van Calster B, McLernon DJ, van Smeden M, et al. Calibration: the Achilles heel of predictive analytics. BMC Med. 2019;17(1):230. doi:10.1186/s12916-019-1466-7

18. Van Calster B, Nieboer D, Vergouwe Y, De Cock B, Pencina MJ, Steyerberg EW. A calibration hierarchy for risk models was defined: from utopia to empirical data. J Clin Epidemiol. 2016;74:167–176. doi:10.1016/j.jclinepi.2015.12.005

19. Steyerberg EW, Vickers AJ, Cook NR, et al. Assessing the Performance of Prediction Models: A Framework for Traditional and Novel Measures. Epidemiology. 2010;21(1):128. doi:10.1097/EDE.0b013e3181c30fb2

20. Transparent Reporting of a Multivariable Prediction Model for Individual Prognosis or Diagnosis (TRIPOD) | Circulation. Accessed June 22, 2025. https://www.ahajournals.org/doi/full/10.1161/CIRCULATIONAHA.114.014508

21. Predicting Outcomes Using the Heart Failure Survival Score in Adults with Moderate or Complex Congenital Heart Disease - Lin - 2015 - Congenital Heart Disease - Wiley Online Library. Accessed June 22, 2025. https://onlinelibrary.wiley.com/doi/abs/10.1111/chd.12229?casa_token=0hDohBZ2Z48AAAAA:x8z9kTsxlKMQ_h0DFHW1HbHahe0jfeopu-PJWSTaBVSDh1_1nrNltUp3YLRb8BQDakw7FiMk407B

22. The Seattle Heart Failure Model | Circulation. Accessed June 22, 2025. https://www.ahajournals.org/doi/full/10.1161/CIRCULATIONAHA.105.584102

23. Selecting patients for heart transplantation: Comparison of the Heart Failure Survival Score (HFSS) and the Seattle Heart Failure Model (SHFM) - ScienceDirect. Accessed June 22, 2025. https://www-sciencedirect-com.proxy.hsl.ucdenver.edu/science/article/pii/S1053249811010059?casa_token=1ojVMNBvRA8AAAAA:EZKx86GKZKe6OgnjwBFK0S00Kz9f-hnc8SXcJPn3wEpgL2mPeman6YIU6DUtLLZO2_B0SA-e

24. Lustig DB, Rodriguez R, Wells PS. Implementation and validation of a risk stratification method at The Ottawa Hospital to guide thromboprophylaxis in ambulatory cancer patients at intermediate-high risk for venous thrombosis. Thromb Res. 2015;136(6):1099–1102. doi:10.1016/j.thromres.2015.08.002

25. Pocock SJ, Ariti CA, McMurray JJV, et al. Predicting survival in heart failure: a risk score based on 39 372 patients from 30 studies. Eur Heart J. 2013;34(19):1404–1413. doi:10.1093/eurheartj/ehs337

26. Development and Validation of a Risk Score Predicting Death Without Transplant in Adult Heart Transplant Candidates | Surgery | JAMA | JAMA Network. Accessed June 22, 2025. https://jamanetwork.com/journals/jama/fullarticle/2814884

27. Murphy RD, Park SY, Allen LA, et al. The Colorado Heart Failure Acuity Risk Model. JACC Adv. 2025;4(1):101449. doi:10.1016/j.jacadv.2024.101449

28. Model for End-stage Liver Disease: Did the New Liver Allocation Policy Affect Waiting List Mortality? | Gastrointestinal Surgery | JAMA Surgery | JAMA Network. Accessed June 22, 2025. https://jamanetwork.com/journals/jamasurgery/fullarticle/400983

29. Evans MD, Diaz J, Adamusiak AM, et al. Predictors of Survival After Liver Transplantation in Patients With the Highest Acuity (MELD ≥40). Ann Surg. 2020;272(3):458. doi:10.1097/SLA.0000000000004211

30. Sadaka F, EthmaneAbouElMaali C, Cytron MA, Fowler K, Javaux VM, O’Brien J. Predicting Mortality of Patients With Sepsis: A Comparison of APACHE II and APACHE III Scoring Systems. J Clin Med Res. 2017;9(11):907–910. doi:10.14740/jocmr3083w

31. Goff RR, Uccellini K, Lindblad K, et al. A change of heart: Preliminary results of the US 2018 adult heart allocation revision. Am J Transplant Off J Am Soc Transplant Am Soc Transpl Surg. 2020;20(10):2781–2790. doi:10.1111/ajt.16010

32. Hanff TC, Harhay MO, Kimmel SE, Birati EY, Acker MA. Update to An Early Investigation of Outcomes with the New 2018 Donor Heart Allocation System in the United States. J Heart Lung Transplant Off Publ Int Soc Heart Transplant. 2020;39(7):725–726. doi:10.1016/j.healun.2020.02.018

33. Trivedi JR, Cheng A, Singh R, Williams ML, Slaughter MS. Survival on the Heart Transplant Waiting List: Impact of Continuous Flow Left Ventricular Assist Device as Bridge to Transplant. Ann Thorac Surg. 2014;98(3):830–834. doi:10.1016/j.athoracsur.2014.05.019

34. Tibrewala A, Chuzi S, Wu T, et al. Impact of Heart Transplant Allocation Change on Waitlist Mortality and Posttransplant Mortality in Patients With Left Ventricular Assist Devices. Circ Heart Fail. 2024;17(11):e011621. doi:10.1161/CIRCHEARTFAILURE.124.011621

35. Association of high-priority exceptions with waitlist mortality among heart transplant candidates - ClinicalKey. Accessed June 22, 2025. https://www-clinicalkey-com.proxy.hsl.ucdenver.edu/#!/content/playContent/1-s2.0-S1053249823018636?returnurl=null&referrer=null

36. Between-center variation in high-priority listing status under the new heart allocation policy - ScienceDirect. Accessed June 22, 2025. https://www-sciencedirect-com.proxy.hsl.ucdenver.edu/science/article/pii/S1600613522088001

